# Design evaluation of a dental implant used in the jawbone D1 – D4 zones

**DOI:** 10.1101/2020.09.21.20199067

**Authors:** Farid Behzadian, Amirhossein Borjali, Mahmoud Chizari

## Abstract

Many people may suffer from missing a tooth or teeth for different reasons. Dental implants are one of the primary solutions which are being used increasingly in the last few decades. In the first stage of the usual procedure, the surgeon uses a drill to create a socket into the jaw bone; then, in the next step, the implant, which is a fine metal screw most of the time, will be inserted into the pre-drilled socket.

This study investigates the biomechanical behaviors of a new dental implant which has a different performance and thread design. The strength of the new implant has been assessed under a daily loading through a series of experimental tests using dummy foam block substitute to the natural human jawbone. The feature of the new design was modified to introduce a new dental implant for brittle jawbones. The study was closely focusing on the design criteria of the implant applicable for D3-D4 zones of the jaw bone.

## Introduction

In order to restore the mechanical function, aesthetics, comfort, and speech of patients with missing a tooth or teeth, dental implants present one of the primary solutions to face this issue [1].

There are a few different types of oral implants at the moment, which some of them are not as popular as it used to be anymore, only in some specific areas. Root-form implants are the type that is being used more often nowadays, and there is a wide range of designs for this type of implants compared to other oral implants [2]. After introducing the Per- Ingvar Branemark titanium screw, the root-form implants became very popular.

Dental implants have advantages such as they do not require grinding of sound teeth (as for fixed bridges) on either side of the missing tooth or teeth, and they look very similar to the natural tooth, so they can be very aesthetic [3]. Moreover, dental implants function as well as natural teeth and can be used to replace multiple teeth with fixed bridges [4]. They also maintain jawbone better than removable dentures [5]; in other words, when there is more than one missing tooth, and they have not been replaced, it can affect the patient’s facial structure by deforming the jaw bone.

However, there are some disadvantages, including dental implants are expensive, and all of them require surgery [6]. Furthermore, they are more difficult for dentists to place than removable partial dentures and require several months for bone integration (most techniques); they might require a specialized dentist to perform the surgery [7].

In a process called Osseointegration, a Root-form implant is screwed to the jawbone in order to play the role of the missing tooth or teeth. However, there is always a chance of failures in using the implants; the patient will be tested for having initial conditions before starting the procedure in order to minimize those chances of failure. There are few essential factors affecting osseointegration, including implant biocompatibility, implant design, implant surface, implant bed, surgical technique and loading condition [8].

It is vital to study and investigate the most popular and common existing designs, and to do so, it was decided to examine numerous companies that are well recognized in the dental implant industry and assess the different products that these companies offer. Five reputable companies, including BioHorizons, Biomet 3i, MEGA’GEN, Straumann, and Zimmer, were chosen and investigated.

In this investigation, it was found that each company has a different method of surface treatment, and almost all of the existing product’s shapes can be categorized into two main groups, including parallel-walled and tapered [9]. However, there are designs that are partially tapered or paralleled walled. As it is evident from their names, the parallel-walled implants have a quite cylindrical shape, and the tapered implants have a conical shape that, in a way, is more similar to the shape of the natural root. For the last couple of years, tapered designs were become more popular comparing to parallel-walled implants [10].

Nearly all of the existing root-form implants have one common feature, which is threaded surface. However, threading in different designs has numerous factors involving Pitch, the number of threads per unit length, and depth between individual threads.

Moreover, there are some other features that different companies use in different designs, such as making a hole through the implant body or subtracting a small part of the implant body known as flute. For example, in the AdVent implant system by Zimmer, there is a hole through the implant body at the bottom section of the implant [11], and in the Branemark implant System by Nobel Biocare in the bottom section of the implant, part of the implant’s body has been subtracted. Flute feature helps the screw to cut through the bone as the implant screws into the socket, and it results in insertion torque and pull-out strength [12].

Generally, the main objective of this study is to design a new dental implant with at list a different feature comparing to existing ones so that it can maximize the rate of success in D3 and D4 bone sites as well as the D1 and D2.

## Methodology

After studying different popular shapes and features, plus how reliable they are, couple concepts were made similar to existing designs, and 3D designs were produced using Siemens NX 7.5 CAD software. 3D models were exported as a specific format, and using a 3D printer called “Dimension 768 Series,” a few numbers of ABS prototypes were made. This specific printer uses a software, Catalyst® EX, that automatically imports STL files, orients the desired part, slices the file, and creates a precise deposition path to build the designing ABS model; however, before this final step, the software makes generate support structures if necessary. The software provides build time, material status, system status information along with management capabilities. One of the many advantages of this particular printer is that it runs unattended and can offer both systems and build status information via e-mail, internet or pager. The printer works on ABS (P400) plastic.

First generated prototypes were used to do some evaluations about the shape and all the features. There was a need to see what the benefits are for each feature according to the application of use, which in this research, one of the objectives was a soft bone application (D3 and D4 bone types). The research key point was to maximize the contact surface between the implant and the surrounding bone, which can improve the permanent stability of the implant and its implantation success rate in D3 and D4 bone sites.

After studying the benefits of each feature and looking thorough research in other potential designs and patents, it was found that MEGA’GEN has a model names MegaFix which is fairly similar to couple existing patents [13] [4,5]. In MegaFix‘s design, the surface has been divided into two sections, and each section has a different method of threading. By considering the corrections in features used in the first prototypes and the idea of having two different sections on the surface, a new 3D design was generated in Siemens NX 7.5. After completing the new design, following the same protocols, and using the same 3D printer, a couple of new ABS samples were created. By looking at the new ABS samples and studying the new shape and features, it was decided to move on to the next stage and make a new sample made of other material than plastic so it can be used for few experiments on dummy foam blocks. Most of the oral implants are made of materials such as Cp titanium, Titanium alloy, Zirconium, and Hydroxyapatite (HA), one type of calcium phosphate ceramic material. However, these materials are quite expensive, so it was decided to make a prototype made of stainless steel, which is a lot cheaper and easier to find. Furthermore, the stainless steel model was made twice the size of the actual dimensions because of the production tools limitations in the workshop.

The stainless steel sample was used to perform a single cycle (pull-out) test on three different foam blocks with three individual densities, including 0.12, 0.16, and 0.24 g/cc (7.5, 10, 15 PCF). The blocks were pre-drilled with a specific depth and diameter. According to the surgical drilling protocols, The diameter of the holes was approximately 1 mm undersized compering to the actual diameter of the design.

After drilling the blocks, a torque screwdriver (Figure 1) was used to measure the torque while screwing the implant into the socket. As a result, it took about 0.65 Nm to screw the implant into the socket on block 0.24g/cc, 0.4 Nm for 0.16 g/cc, and 0.2 Nm for 0.12g/cc. A servo-hydraulic machine, Instron 8501 (Figure 2), was used to carry the pull-out test, which is capable of recording tensile and compress loads at different loading rates [14, 15]. The machine required to be initialized with some initial information including test type, the diameter of the drilled hole, the screw’s diameter, torque setting, experiment site’s temperature, experiment site’s humidity, and speed rate.

**Figure 1:**
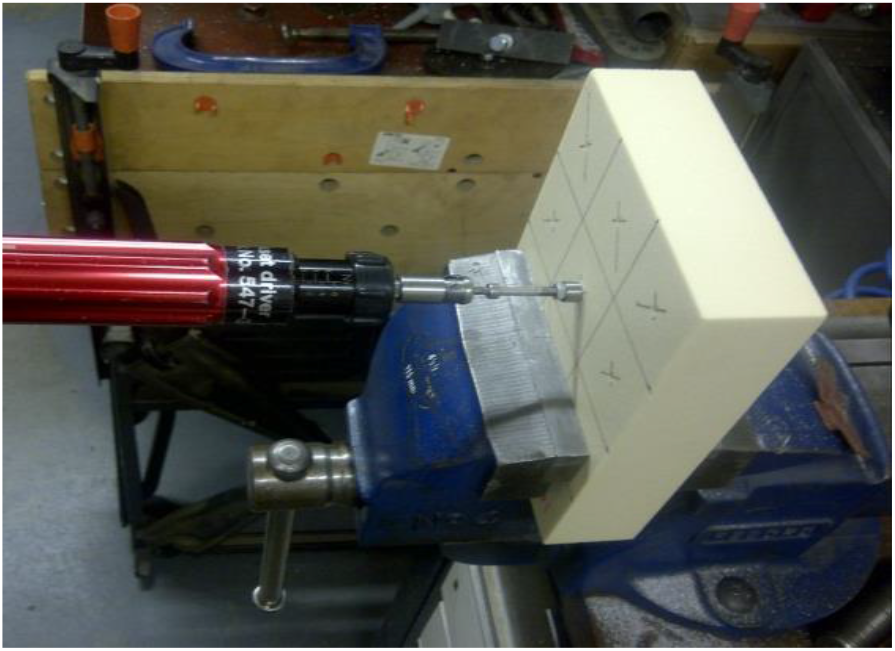
The torque required to insert the new implant into the foam block was measured using a torque meter.

**Figure 2:**
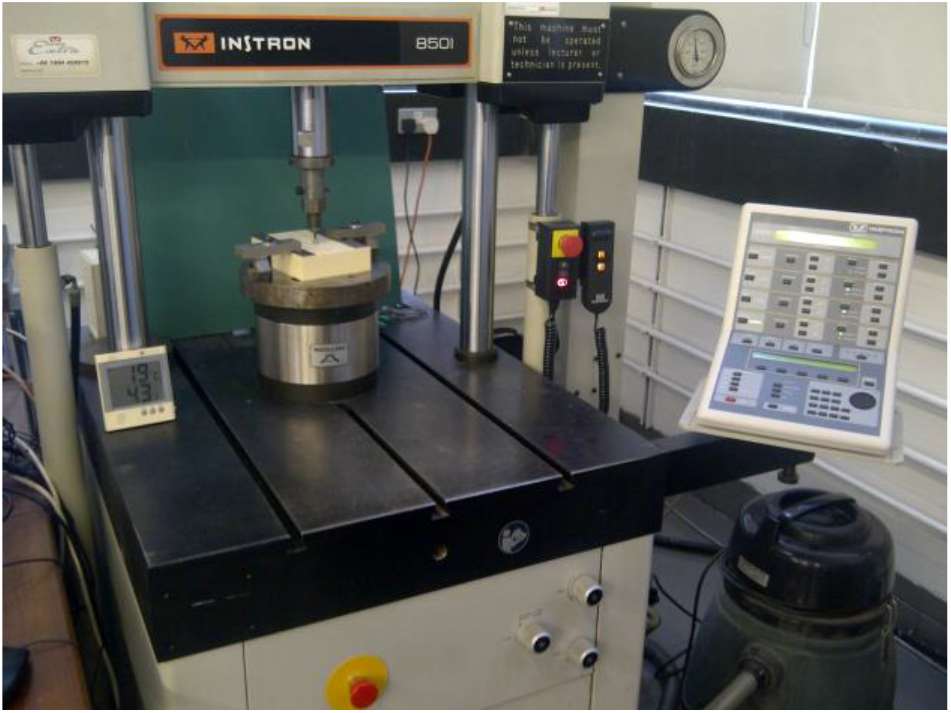
Performing pull-out test on a foam block using an Instron 8501 testing machine.

## Results and Discussion

Parallel-walled and tapered implants both can provide primary implant stability in soft bone and hard bone depending on the design. Regarding the implant design, the surgeon must know in advance the bone density plus the anatomical complexities of the surgical site; then, according to this knowledge, an implant design can be chosen and placed.

As was mentioned earlier, parallel-walled and tapered both can be used in soft or hard bone, but there are individual sites that one of these two types is more advised. The best choice to place in dense D1 bone (anterior segment of the atrophic, edentulous mandible) is using parallel cylinder implants to avoid over-compression of the dense bone, causing impairment of blood supply. On the other hand, To get the best primary stability for placing in the soft bone such as D3 (posterior maxilla), it is more logical and suitable to place conical tapered implants. However, according to some previous work, cylindrical screws have constant insertion torque in any given material as long as the pitch is held constant [16]. This statement also applies and holds true for the cylindrical portion of a combined design. In comparison, it was also found that the placement torque of tapered screws increases throughout placement [10]; by considering this, the insertion torque can be a very critical factor when it comes to tapered designs for D3 and D4 bone sites.

In surgical procedures, one of the parameters that always needs to be measured is the torque that the sergeant uses to screw the implant into the socket. The insertion torque should be of a reasonable level because strong insertion torques may result in stress absorptions around the implant, with consequent bone resorption.

As a result, it was essential to check if the insertion torque is a moderate level. As was mentioned earlier, the torque was measured for each block 0.12, 0.16, 0.24g/cc was 0.2, 0.4, 0.65 Nm, respectively, which is just about the torque average controls (14.80+/-1 N cm and 66.31+/-0.9) (P<0.05) [17] (Figure 3).

**Figure 3:**
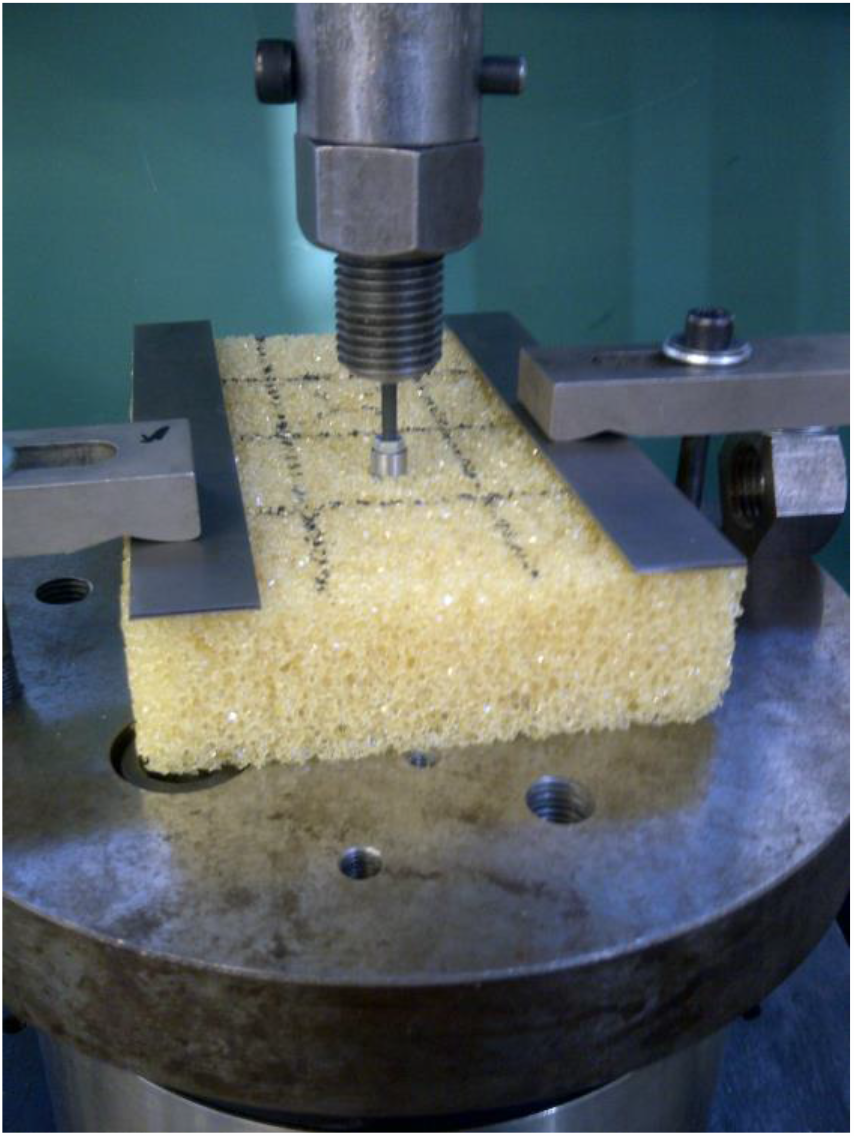
The implant has initialized in the artificial bone bock with a density of 0.12 g/cc and ready to start the pull-out experiment.

The Benefit of creating a hole through the implant’s body is as bone tissue can penetrate into the implant’s body through these holes, the contact area with the bone is improved, and therefore, a high bonding force can be gained. Moreover, the reason for having a small section subtracted from each body is to get better abutment stability. Furthermore, these holes can sometimes act as flutes depending on their positions and angles on the implant’s body.

The final design in this study has a unique feature, which was a different surface shape rather than thread. So as it was mentioned earlier, the surface of the implant was divided into two sections, including upper and lower section. The upper section has the threaded design, and the lower section, which enters the socket first, has that new unique surface shape. This new surface shape benefits in maximizing the contact area between the implant surface and the bone surrounding it. The lower section of the implant can also be used for some surface treatment or coating even to maximize the contact surface between the implant and the bone more beneficial. Note that the surface coating can be done on threaded surfaces, although there is a risk of coating damage due to high insertion force.

In Table 1, the average results for each block have been shown. The jaw bone has a natural property similar to a sponge, and as the density of bone decreases, it becomes more brittle, and the number of empty spaces inside the property increases. According to Table 1, as it was expected, the maximum load has been increased by growing the density of blocks, and that is because when the machine tries to pull out the screw, as the density increases, it takes more load to pull out the screw. Furthermore, it is obvious that the tensile stress values at maximum load have been increased as the bone density increases. From Table 1, it is shown that the average load and the average tensile stress increases directly with the various block densities shown in the first column.

**Table 1:** Reading results for a series of experimental tests using the newly designed dental implant and dummy foam blocks.

| Block density (gr/cc) | Load (N) | Extension (mm) | Tensile stress (MPa) | Tensile strain (mm) |
| --- | --- | --- | --- | --- |
| 0.12 | 209.1 | 0.4 | 3.29 | 0.036 |
| 0.16 | 297.1 | 0.3 | 4.67 | 0.030 |
| 0.24 | 332.5 | 0.3 | 5.23 | 0.026 |

## Conclusion

In this study, the theoretical background research, as well as the experimental results due to the experiments, showed that the new proposed design, this model can provide all mechanical needs as a suitable root form implant, and it has the potential to be a solution for success rate improvement in implantation in soft bone sites as well as hard bone sites. This study also verifies that there is a possibility to find and work on different surface shapes rather that thread for specific applications to accomplish better results.

## Data Availability

The data have not been stored in any data repositories.

## References

[1] N. Garg and A. Garg, Textbook of operative dentistry: Boydell & Brewer Ltd, 2010.

[2] L. Gaviria, J. P. Salcido, T. Guda, and J. L. Ong, “Current trends in dental implants,” Journal of the Korean Association of Oral and Maxillofacial Surgeons, vol. 40, pp. 50–60, 2014.

[3] R. Rubbert, “Customized dental prosthesis for periodontal-or osseointegration, and related systems and methods,” ed: Google Patents, 2010.

[4] J. A. Bassett, M. Collins, and S. Cahill, “Patient-specific implants with improved osseointegration,” ed: Google Patents, 2017.

[5] C. Ravi Kumar, K. Pratap, and G. Venkateswararao, “Dental Implants As An Option In Replacing Missing Teeth: A Patient Awareness Survey In Khammam, Andhra Pradesh,” Indian Journal of Dental Sciences, vol. 3, 2011.

[6] S. Mathuriya and S. Agarwal, “To study awareness of patients about implant and willingness for implant,” J Appl Dent Med Sci, vol. 1, pp. 10–8, 2015.

[7] Y. Oshida, E. B. Tuna, O. Aktören, and K. Gençay, “Dental implant systems,” International journal of molecular sciences, vol. 11, pp. 1580–1678, 2010.

[8] P. C. Chang, N. P. Lang, and W. V. Giannobile, “Evaluation of functional dynamics during osseointegration and regeneration associated with oral implants,” Clinical oral implants research, vol. 21, pp. 1–12, 2010.

[9] M. Alshehri and F. Alshehri, “Influence of implant shape (tapered vs cylindrical) on the survival of dental implants placed in the posterior maxilla: a systematic review,” Implant Dentistry, vol. 25, pp. 855–860, 2016.

[10] M. V. dos Santos, C. N. Elias, and J. H. Cavalcanti Lima, “The effects of superficial roughness and design on the primary stability of dental implants,” Clinical implant dentistry and related research, vol. 13, pp. 215–223, 2011.

[11] A. Warreth, E. McAleese, P. McDonnell, R. Slami, and M. S. Guray, “Dental implants and single implant-supported restorations,” 2013.

[12] R. Chowdhary, On efficacy of implant thread design for bone stimulation: Malmö University, Faculty of Odontology, 2014.

[13] W. Y. Hung, “Dental implant,” ed: Google Patents, 2011.

[14] A. Avanzini, “Effect of cyclic strain on the mechanical behavior of virgin ultra-high molecular weight polyethylene,” Journal of the mechanical behavior of biomedical materials, vol. 4, pp. 1242–1256, 2011.

[15] A. Avanzini and D. Gallina, “Effect of cyclic strain on the mechanical behavior of a thermoplastic polyurethane,” Journal of engineering materials and technology, vol. 133, 2011.

[16] S.-W. Wu, C.-C. Lee, P.-Y. Fu, and S.-C. Lin, “The effects of flute shape and thread profile on the insertion torque and primary stability of dental implants,” Medical engineering & physics, vol. 34, pp. 797–805, 2012.

[17] K. Akça, A. Kökat, A. Cömert, M. Akkocaoğlu, I. Tekdemir, and M. Cehreli, “Torque-fitting and resonance frequency analyses of implants in conventional sockets versus controlled bone defects in vitro,” International journal of oral and maxillofacial surgery, vol. 39, pp. 169–173, 2010.

